# More complaints than findings - Long-term pulmonary function in children and adolescents after COVID-19

**DOI:** 10.1101/2021.06.22.21259273

**Authors:** Leona Knoke, Anne Schlegtendal, Christoph Maier, Lynn Eitner, Thomas Lücke, Folke Brinkmann

**Author notes:** Contributed equally as first authors (Leona Knoke, Anne Schlegtendal). This work is part of the thesis of Leona Knoke. **Corresponding author** Folke Brinkmann, Department of Paediatric Pneumology, University Children’s Hospital, Ruhr-University Bochum Alexandrinenstraße 5, 44791 Bochum, Germany. **Contributors** CM, TL, FB developed the study design and protocol and supervised patient management. LK drafted the manuscript with contributions of intellectual content from AS, LE, CM, TL and FB. LK, AS, LE collected the data. LK, AS, CM and FB examined the data and undertook statistical analyses. All authors have seen and approved the final version of the manuscript. FB is guarantor and attests that all named authors and contributors meet authorship criteria and that no others meeting such criteria have been omitted. **Clinical trial ID** DRKS00022434, German clinical trial register. **Competing interests** None declared. **Patient consent for publication** Not required. **Ethics approval** Ethical approval was provided by the Ethics Committee of the Ruhr-University Bochum, Germany (No. 20-6927). **Checklist for the appropriate study type** STROBE and RECORD checklists are included as supplementary appendices. **Data availability statement** Data are available upon reasonable request.

## Abstract

**Background:** The frequency of persistent symptoms after coronavirus disease 2019 (COVID-19) in adults varies from 4.5% to 87%. Pulmonary function can also show long-term impairment in adults: 10% of hospitalised adults had reduced spirometry values, and 24% had decreased diffusion capacity. To date, only preliminary evidence is available on persistent respiratory sequelae in children and adolescents, therefore our objective was to examine the long-term effects of COVID-19 on pulmonary function in this age group.

**Methods:** Multiple-breath washout, body plethysmography, and diffusion capacity testing were performed after an average of 2.6 months (range 0.4–6.0) following COVID-19 in 73 children and adolescents (age 5–18 years) with different disease severity. Cases were compared to 45 controls with and without infection within six months prior to assessment after exclusion of severe acute respiratory coronavirus-2 infection (SARS-CoV-2).

**Results:** Of the 19 patients (27.1%) who complained about persistent or newly emerged symptoms since COVID-19, 8 (11.4%) reported respiratory symptoms. Comparing patients with COVID-19 to controls, no significant differences were detected in frequency of abnormal pulmonary function (COVID-19: 12, 16.4%; controls: 12, 27.7%; OR 0.54, 95% CI 0.22–1.34). Only two patients with persistent respiratory symptoms showed abnormal pulmonary function. Multivariate analysis revealed reduced forced vital capacity (*p*=0.045) in patients with severe infection regardless of SARS-CoV-2 infection.

**Discussion:** Pulmonary function is rarely impaired in children and adolescents after COVID-19, except of those with severe infection. The discrepancy between persistent respiratory symptoms and normal pulmonary function suggests a different underlying pathology such as dysfunctional breathing.

## INTRODUCTION

The clinical course of children and adolescents with SARS-CoV-2 infection is mostly less severe than in adults [1].

Long-COVID sums up all symptoms that persist or newly emerge four weeks to beyond twelve weeks after acute SARS-CoV-2 infection [2].

In one study, up to 87% of adults hospitalised due to COVID-19 infection reported persistent symptoms (for example fatigue, dyspnoea, cough, gastrointestinal symptoms) about two months after COVID-19 infection [3]. In another cohort with in- and outpatients in the same age range, only 4.5% were still symptomatic with similar symptoms after eight weeks [1]. There is only preliminary evidence of persistent symptoms in children: Buonsenso et al. described that 52.7% of their study population reported at least one persistent symptom equal to or more than four months after SARS-CoV-2 infection, while 14.7% stated they suffered persistent respiratory problems [4].

Long-lasting deterioration in pulmonary function has been described in adults [5–7]. Lerum et al. reported that about 10% of adults hospitalised with COVID-19 had reduced values for forced vital capacity (FVC) and forced expiratory volume in one second (FEV_1_), and 24% showed significantly reduced diffusion capacity at a three-month follow-up visit [5]. However, frequency and extent of long-term changes in pulmonary function in children and adolescents following COVID-19 infection are barely examined. Bottino et al performed spirometry and diffusion capacity testing in seven children about two months after recovery from mild SARS-CoV-2 infection and did not find any abnormalities [8].

To evaluate the impact of respiratory sequelae in children is important given the fact that most children worldwide will potentially get infected with SARS-CoV-2 as long as vaccines are predominantly reserved for adults and high-risk groups.

We analysed pulmonary function of children and adolescents after COVID-19 and compared their results to a group of controls after exclusion of SARS-CoV-2 infection.

## PATIENTS AND METHODS

### Study design and population

We conducted a single-centre, cross-sectional, prospective study to assess pulmonary function after COVID-19 in children and adolescents from 5 to 18 years of age. This study was part of a still ongoing study at the University Children’s Hospital of Bochum, Germany, assessing rates of SARS-CoV-2 seroconversion in children and adolescents in western Germany (CorKid).

We recruited from August 2020 to March 2021 and included subjects with positive SARS-CoV-2 antibodies tested in the CorKid study, as well as in- and outpatients who tested positive for SARS-CoV-2 using polymerase chain reaction (PCR) at the University Children’s Hospital of Bochum or in an outpatient practice in the region. Children with negative antibodies for SARS-CoV-2 and no other evidence of SARS-CoV-2 infection from the CorKid study served as controls. Exclusion criteria were inability to perform lung function testing and missing written consent.

The subjects and/or their guardians answered questionnaires regarding their medical history, especially acute and chronical pulmonary diseases, as well as current and previous medications. Any infections within six months prior to the assessment and their date, duration and symptoms, as well as PCR test results for SARS-CoV-2 were recorded. We also documented persistent symptoms after COVID-19 infection in the COVID-19 group.

According to these data, we subdivided the COVID-19 group into those who were symptomatic and those who were asymptomatic, and showed signs of respiratory tract infections (if cough, rhinitis, sore throat and/or dyspnoea was confirmed) and other types. Severity of infection was subclassified into severe and non-severe. An infection was considered severe if it included dyspnoea, fever >38.5°C for >5 days, bronchitis or pneumoniae, and/or hospitalisation >1 day.

Patients were classified as having chronic lung disease if they had a known medically diagnosed pulmonary disease like bronchial asthma or if they had one of the following: recurrent wheeze, pneumonia in the year before measurement, previous or current use of inhaled steroids for at least 4 weeks or long-lasting productive cough (≥8 weeks).

We also divided the subjects into two groups (follow-up 0–3 and 4–6 months) depending on the interval between infection and assessment.

### Pulmonary function testing

Body plethysmography and diffusing capacity testing for carbon monoxide (CO) were performed using MasterScreen Body/Diff (Vyaire, Hoechberg, Germany) according to American Thoracic Society/European Respiratory Society guidelines [9, 10]. We measured FEV_1_, FVC, mean expiratory flow at 75% (MEF_75_) and MEF_25_, as well as diffusing capacity of the lung for carbon monoxide (DLCO) and diffusing capacity divided by the alveolar volume (DLCO/VA) standardised to *Z*-scores using Global Lung Initiative reference values [11, 12]. DLCO and DLCO/VA were adjusted for haemoglobin levels. N_2_ multiple-breath washout was measured using Exhalyzer D (EcoMedics AG, Duernten, Switzerland) according to current guidelines [13]. We reported the Lung Clearance Index at 2.5% of starting concentration (LCI_2.5%_) as recommended.

All pulmonary function tests were done by specially trained staff and assessed by two paediatric pneumologists blinded to the patient’s SARS-CoV-2 status.

Pulmonary function was defined as abnormal, if at least one measured parameter was pathological (LCI_2.5%_ >7.9, FVC *Z*-score, FEV_1_ *Z*-score, MEF_75_ *Z*-score, MEF_25_ *Z*-score, DLCO *Z*-score, DLCO/VA *Z*-score <-1.96) or there was at least one borderline parameter (LCI_2.5%_ >7.0, FVC *Z*-score, FEV_1_ *Z*-score, MEF_75_ *Z*-score, MEF_25_ *Z*-score, DLCO *Z*-score, DLCO/VA *Z*-score <–1 ≥–1.96) for two different pulmonary function tests (N_2_ multiple-breath washout, body plethysmography, diffusion capacity).

Imaging of the lungs was performed only when clinically necessary.

### Statistics

Descriptive statistics: We calculated mean, median, range and standard deviation for continuous variables. Categorical values were expressed as absolute values and percentages.

Inferential statistics: Odds Ratios (ORs) and 95% confidence intervals (95% CIs) were calculated for categorical values. Normal distribution was proven for all pulmonary function parameters either using the Komolgorov-Smirnov test or the Levene test. Independent *t*-test or analysis of variance (ANOVA) was used for between-group comparisons. Post-hoc analysis was performed using least significant difference. All statistical analysis were performed using SPSS version 27 (SPSS for Windows, SPSS Inc., Chicago, IL, USA). A *p*-value of 0.05 or less was considered statistically significant.

### Ethical approval

The ethics committee of the Ruhr-University, Bochum, Germany, approved the project (register number: 20-6927). All participants and/or their parents were informed about the study and provided written informed consent.

## RESULTS

We enrolled 73 subjects with confirmed SARS-CoV-2 infection, 26 (35.6%) of whom were symptomatic during the acute phase. Of the 45 subjects who served as controls, 14 (31.1%) had any symptomatic infection other than COVID-19 within six months before the assessment. Excluding six COVID-19 patients without PCR, the mean interval between infection and date of testing was similar in both groups (COVID-19: 2.59 months (range 0.4–6.0); controls: 3.43 months (range 1.03–6.3)).

The epidemiological data were similar in both groups (see Table 1a).

**Table 1a:** Epidemiological data.

| Epidemiological data | COVID-19 patients |  |  | Controls |  |  | OR (95% CI)<sup>‡</sup> |
| --- | --- | --- | --- | --- | --- | --- | --- |
|  | All | Symptomatic infection within the past 6 months | Asymptomatic infection within the past 6 months | All | Any other infection within the past 6 months | No infection within the past 6 months |  |
| **N** | 73 | 27* | 46 | 45 | 14 | 31 |  |
| **Sex, female** | 38 (52.0) | 14 (51.9) | 24 (52.2) | 28 (62.2) | 11 (78.6) | 17 (54.8) | 0.66 (0.31–1.41) |
| **Age, years, mean ± SD** | 10.82 (± 3.25) | 10.48 (± 3.19) | 11.02 (± 3.3) | 10 (± 3.5) | 9.14 (± 3.39) | 10.39 (± 3.53) |  |
| Age group, 5–8 years | 21 (28.8) | 7 (25.9) | 14 (30.4) | 18 (40) | 8 (57.1) | 10 (32.3) | 0.61 (0.28–1.32) |
| Age group, 9–12 years | 29 (39.7) | 13 (48.2) | 16 (34.8) | 16 (35.6) | 2 (14.3) | 14 (45.2) | 1.19 (0.55–2.58) |
| Age group, 13–18 years | 23 (31.5) | 7 (25.9) | 16 (34.8) | 11 (24.4) | 4 (28.6) | 7 (22.6) | 1.42 (0.61–3.29) |
| **Gestational age, premature** | 4 (5.6)<sup>2</sup> | 2 (8)<sup>2</sup> | 2 (4.4) | 5 (11.1) | 2 (14.3) | 3 (9.7) | 0.48 (0.12–1.88) |
| **Neonatal intensive care unit** | 13 (18.3)<sup>2</sup> | 8 (32)<sup>2</sup> | 5 (10.9) | 4 (8.9) | 2 (14.3) | 2 (6.5) | 2.3 (0.7–7.55) |
| With oxygen supplementation | 4 (5.6)<sup>2</sup> | 2 (8)<sup>2</sup> | 2 (4.4) | 1 (2.2) | 0 | 1 (3.2) | 2.63 (0.28–24.29) |
| **Pulmonary disease, previous or current** | 17 (23.3) | 8 (29.6) | 9 (19.6) | 10 (22.2) | 3 (21.4) | 7 (22.6) | 1.06 (0.44–2.58) |

SD, standard deviation; OR, odds ratio; CI, confidence interval
Data are expressed as *n* (%), unless specified otherwise
\* For one patient, the documented symptomatic infection within six months prior to the assessment was probably not SARS-CoV-2 infection
<sup>1</sup> e.g., cough or dyspnoea; <sup>2</sup> missing values *n* = 2, ‡ OR and 95% CI calculated for all COVID-19 vs. all controls

Disease severity and type of infection did not show significant differences between the two groups. However, 70.4% (*n* = 19) of children and adolescents with symptomatic SARS-CoV-2 infection did not have respiratory symptoms, whereas 92.9% (*n* = 13) of infections in the control group involved the respiratory tract. Rhinitis and cough were significant more often in the control group than in the COVID-19 group (rhinitis: OR 0.14, 95% CI 0.03–0.62; cough: OR 0.24, 95% CI 0.06–0.95). Other assessed symptoms did not show significance between the two groups (see Table 1b for more information).

**Table 1b:**
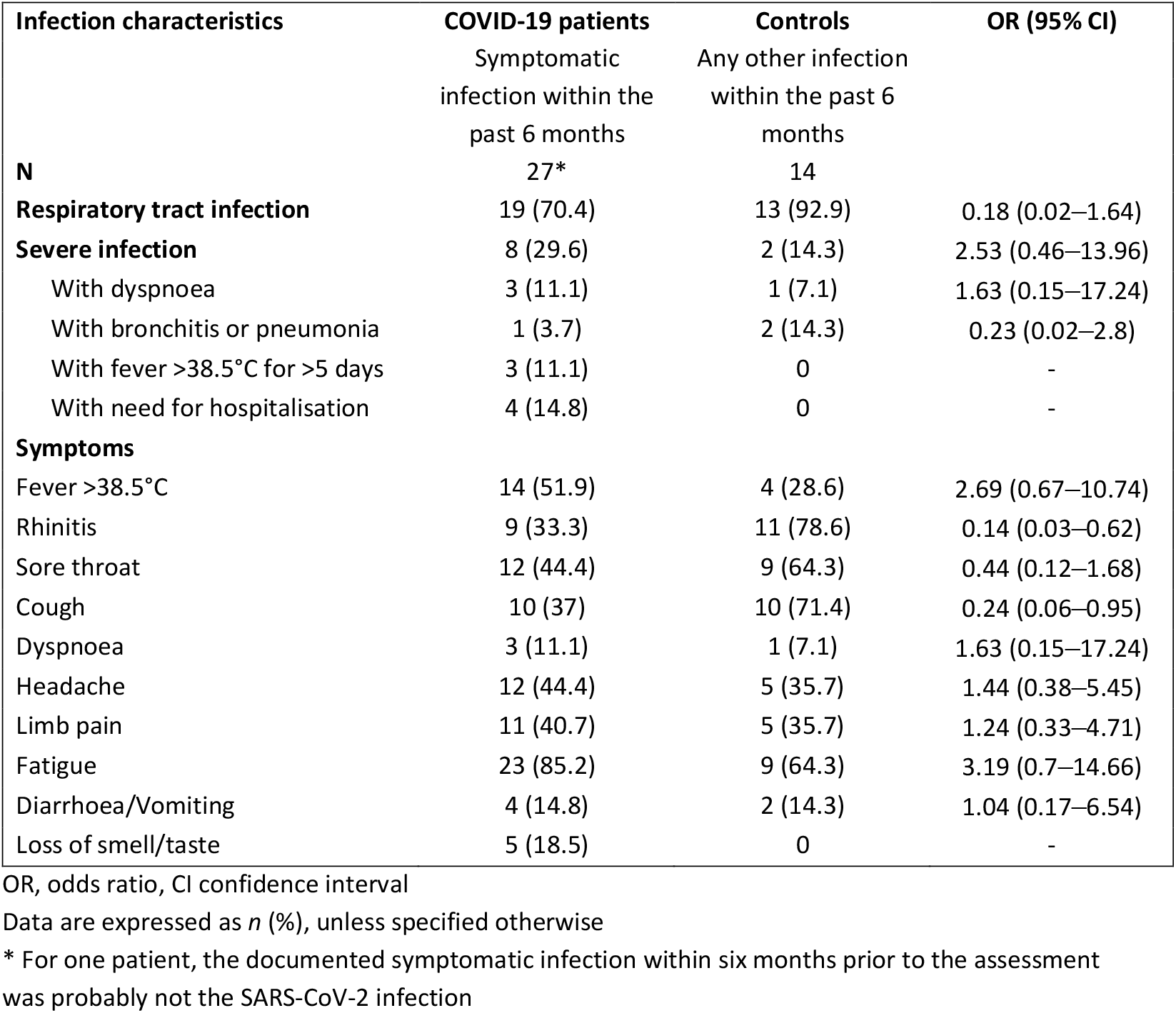
Infection characteristics.

### Long-term complaints

Nineteen (27.1%, data were only available for 70 subjects) children and adolescents of the COVID-19 group reported persistent or newly emerged symptoms after SARS-CoV-2 infection. Eight (11.4%) reported at least one respiratory symptom, six (8.6%) of whom suffered ongoing breathing problems and two (2.9%) persistent cough. The mean age of children and adolescents with respiratory long-term symptoms was similar to all patients with SARS-CoV-2 infection (all COVID-19: 10.82 ± 3.25, patients with respiratory long-term symptoms: 11.75 ± 3.92 [mean ± standard deviation in years]). In two patients, these respiratory problems newly emerged after asymptomatic SARS-CoV-2 infection. Ten (14.3%) suffered from fatigue; five patients (7.1%) had both persistent respiratory symptoms and fatigue syndrome.

### Pulmonary function testing

Comparing the two groups of children and adolescents with and without SARS-CoV-2 infection, there were no significant differences in LCI_2.5%_ (absolute) and age-related *Z*-scores of FVC, FEV_1_, MEF_75_, MEF_25,_ DLCO and DLCO/VA. Details are shown in Table 2 and Figure 1.

**Table 2:** Pulmonary function parameters.

| Pulmonary function parameters | COVID-19 patients |  |  | Controls |  |  |
| --- | --- | --- | --- | --- | --- | --- |
|  | All | Symptomatic infection within the past 6 months | Asymptomatic infection within the past 6 months | All | Any other infection within the past 6 months | No infection within the past 6 months |
| **N<sub>2</sub> Multiple-breath washout** |  |  |  |  |  |  |
| N | 68 | 27 | 41 | 40 | 11 | 29 |
| LCI<sub>2.5</sub>, absolute | 6.75 ± 0.73 | 6.79 ± 0.62 | 6.72 ± 0.81 | 6.88 ± 0.61 | 6.79 ± 0.47 | 6.91 ± 0.66 |
| Abnormal values | 3 (4.4) | 1 (3.7) | 2 (4.9) | 3 (7.5) | 0 | 3 (10.3) |
| **Body plethysmography** |  |  |  |  |  |  |
| N | 71 | 26 | 45 | 45 | 14 | 31 |
| FVC, Z-score | −0.21 ± 1.06 | −0.26 ± 1.04 | −0.19 ± 1.08 | −0.21 ± 1.22 | −0.25 ± 1.53 | −0.19 ± 1.08 |
| Abnormal values | 5 (7) | 1 (3.9) | 4 (8.9) | 5 (11.1) | 2 (14.3) | 3 (9.7) |
| FEV<sub>1</sub>, Z-score | 0.3 ± 1.04 | 0.34 ± 1.04 | 0.28 ± 1.05 | 0.27 ± 1.19 | 0.2 ± 1.34 | 0.3 ± 1.13 |
| Abnormal values | 0 | 0 | 0 | 1 (2.2) | 1 (7.1) | 0 |
| MEF<sub>75</sub>, Z-score | 0.28 ± 1.1 | −0.03 ± 1.13 | 0.46 ± 1.05 | 0.12 ± 1.09 | −0.35 ± 1.38 | 0.34 ± 0.87 |
| Abnormal values | 3 (4.2) | 2 (7.7) | 1 (2.2) | 1 (2.2) | 1 (7.1) | 0 |
| MEF<sub>25</sub>, Z-score | 0.86 ± 0.94 | 0.91 ± 0.68 | 0.83 ± 1.07 | 1.19 ± 1 | 1.04 ± 0.88 | 1.26 ± 1.06 |
| Abnormal values | 0 | 0 | 0 | 0 | 0 | 0 |
| **Diffusion capacity** |  |  |  |  |  |  |
| N | 38 | 10 | 28 | 27 | 4 | 23 |
| DLCO, Z-score | 2.03 ± 2.35 | 2.56 ± 2.27 | 1.84 ± 2.39 | 1.55 ± 1.91 | 2.74 ± 2.7 | 1.35 ± 1.74 |
| Abnormal values | 2 (5.3) | 0 | 2 (7.1) | 1 (3.7) | 0 | 1 (4.4) |
| DLCO/VA, Z-score | 0.02 ± 0.76 | −0.11 ± 1.09 | 0.07 ± 0.62 | −0.21 ± 0.94 | 0.05 ± 1.39 | −0.25 ± 0.88 |
| Abnormal values | 0 | 0 | 0 | 0 | 0 | 0 |

LCI<sub>2.5</sub>, Lung Clearance Index at 2.5% of starting concentration; FVC, Forced Vital Capacity; FEV<sub>1</sub>, Forced Expiratory Volume in the first Second; MEF, Mean Expiratory Flows at different pulmonary volume levels (75, 25); DLCO, Diffusing Capacity of the lungs for Carbon Monoxide; VA, Alveolar Volume All pulmonary function parameters are presented as mean ± standard deviation. Abnormal values are presented in *n* (%).

**Figure 1:**
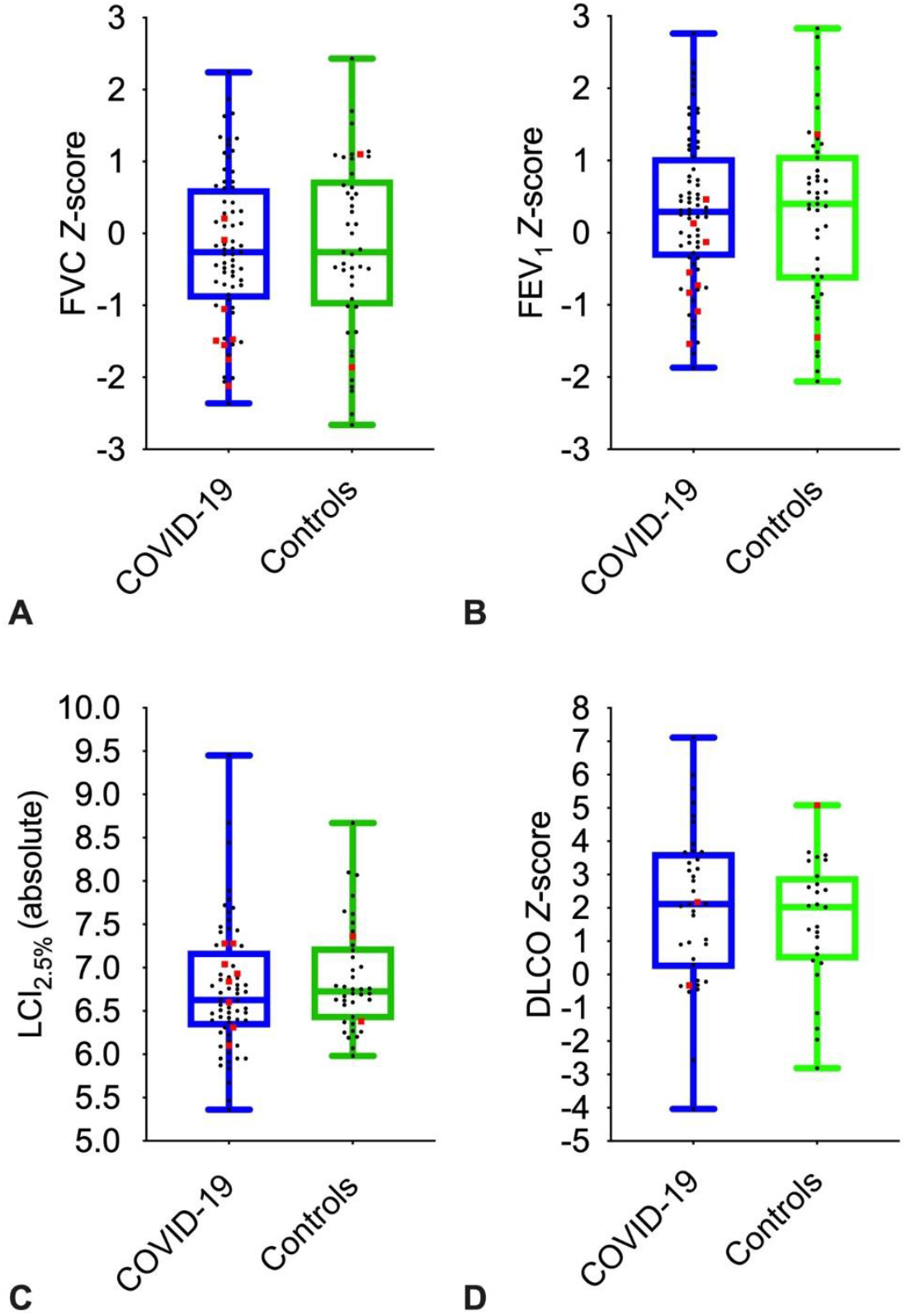
Depiction of pulmonary function parameters for COVID-19 and controls. Boxplots show medians, quartiles, minimum and maximum values. The dots represent the individual values of participants. Participants with severe infection within the last six months are marked as red squares. LCI_2.5%_, Lung Clearance Index at 2.5% of starting concentration; FVC, Forced Vital Capacity; FEV_1_, Forced Expiratory Volume in the first Second; DLCO, Diffusion Capacity of the lungs for Carbon Monoxide

Subjects suffering from severe infection within six months prior to the assessment had significant reduction in FVC (*p* = 0.045) and MEF_75_ (*p* = 0.002) compared to those with non-severe infection and asymptomatic infection. Neither site of infection nor COVID-19 status influenced pulmonary function (see Figure 2). MEF_25_, DLCO, DLCO/VA and LCI_2.5%_ did not show any significant differences between children and adolescents with severe, non-severe or asymptomatic infection six months prior to the assessment.

**Figure 2:**
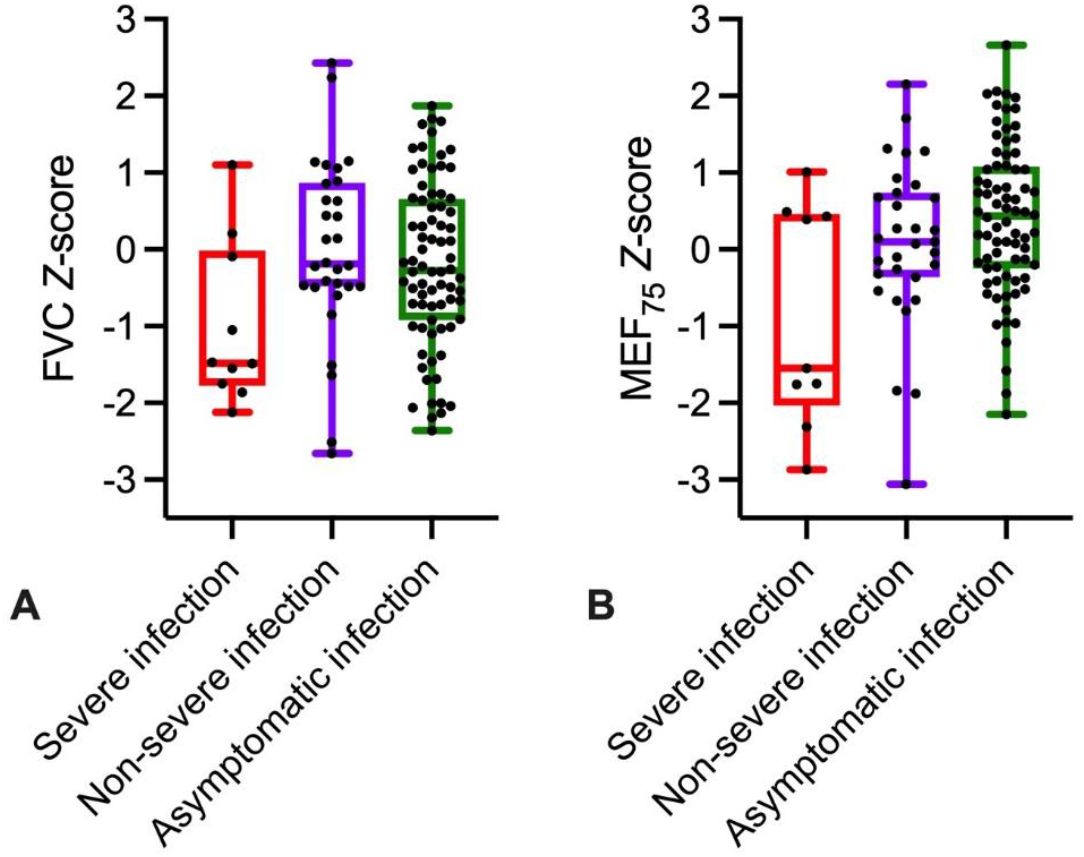
Depiction of **A** FVC *Z*-score and **B** MEF_75_ *Z*-score of all participants (COVID-19 and controls) divided into severe, non-severe and asymptomatic infection, respectively no infection in the control group. Boxplots show medians, quartiles, minimum and maximum values. The dots represent the individual values of participants. FVC, Forced Vital Capacity; MEF_75_, Mean Expiratory Flow at 75%

Overall, 16.4% (*n* = 12) of the children and adolescents after COVID-19 infection had abnormal pulmonary function compared to 27.7% (*n* = 12) without SARS-CoV-2 infection (OR 0.54, 95% CI 0.22–1.34). Comparing all subjects with and without abnormal pulmonary function, no influence of SARS-CoV-2 status, previous or current pulmonary disease, symptomatic infection or severity of infection was found. Most abnormal pulmonary functions occurred in the 5–8-years age group (see Table 3). This effect was mainly visible in the measurements of LCI_2.5%_ and FVC (see Figure 3). Two of the eight patients reporting persistent respiratory complaints, and three of the ten suffering fatigue syndrome showed abnormal pulmonary function.

**Table 3:** Characteristics of participants with and without abnormal^1^ pulmonary function.

| Participant characteristic | Abnormal pulmonary function | Normal pulmonary function | OR (95% CI) |
| --- | --- | --- | --- |
| **N** | 24 | 94 |  |
| **Positive SARS-CoV-2 status** | 12 (50) | 61 (64.9) | 0.54 (0.22–1.34) |
| **Sex, female** | 10 (41.7) | 56 (59.6) | 0.48 (0.2–1.2) |
| **Age, years, mean ± SD** | 8.63 (± 3.7) | 10.99 (± 3.1) | |
| Age group, 5–8 years | 17 (70.8) | 22 (23.4) | 7.95 (2.92–21.63) |
| Age group, 9–12 years | 3 (12.5) | 42 (44.7) | 0.18 (0.05–0.63) |
| Age group, 13–18 years | 4 (16.7) | 30 (31.9) | 0.43 (0.13–1.36) |
| **Gestational age, premature** | 3 (12.5) | 6 (6.4) | 2.1 (0.48–9.07) |
| **Neonatal intensive care unit** | 3 (12.5) | 14 (14.9) | 0.82 (0.21–3.11) |
| With oxygen supplementation | 2 (8.3) | 3 (3.2) | 2.76 (0.43–17.52) |
| **Pulmonary disease, previous or current** | 6 (25) | 21 (22.3) | 1.16 (0.41–3.29) |
| **Infection within 6 months prior to assessment** | 9 (37.5) | 32 (34) | 1.16 (0.46–2.95) |
| With severe infection | 4 (16.7) | 6 (6.4) | 2.93 (0.76–11.37) |
| With hospitalisation | 1 (4.2) | 3 (3.2) | 1.32 (0.13–13.27) |

SD, standard deviation; OR, odds ratio; CI, confidence interval
Data are expressed as n (%), unless specified otherwise
<sup>1</sup> Abnormal pulmonary function defined as at least one measured parameter is pathological (LCl<sub>2.5%</sub> >7.9, FVC Z-score, FEV<sub>1</sub> Z-score, MEF<sub>75</sub> Z-score, MEF<sub>25</sub> Z-score, DLCO Z-score, DLCO/VA Z-score <–1.96) or at least one borderline parameter (LCl<sub>2.5%</sub> >7.0, FVC Z-score, FEV<sub>1</sub> Z-score, MEF<sub>75</sub> Z-score, MEF<sub>25</sub> Z-score, DLCO Z-score, DLCO/VA Z-score <–1 ≥ –1.96) in two different pulmonary function tests (N<sub>2</sub> multiple-breath washout, body plethysmography, diffusion capacity).
LCl<sub>2.5%</sub>, Lung Clearance Index at 2.5% of starting concentration; FVC, Forced Vital Capacity; FEV<sub>1</sub>, Forced Expiratory Volume in the first Second; MEF, Mean Expiratory Flows at different pulmonary volume levels (75, 25); DLCO, Diffusing Capacity of the lungs for Carbon Monoxide; VA, Alveolar Volume

**Figure 3:**
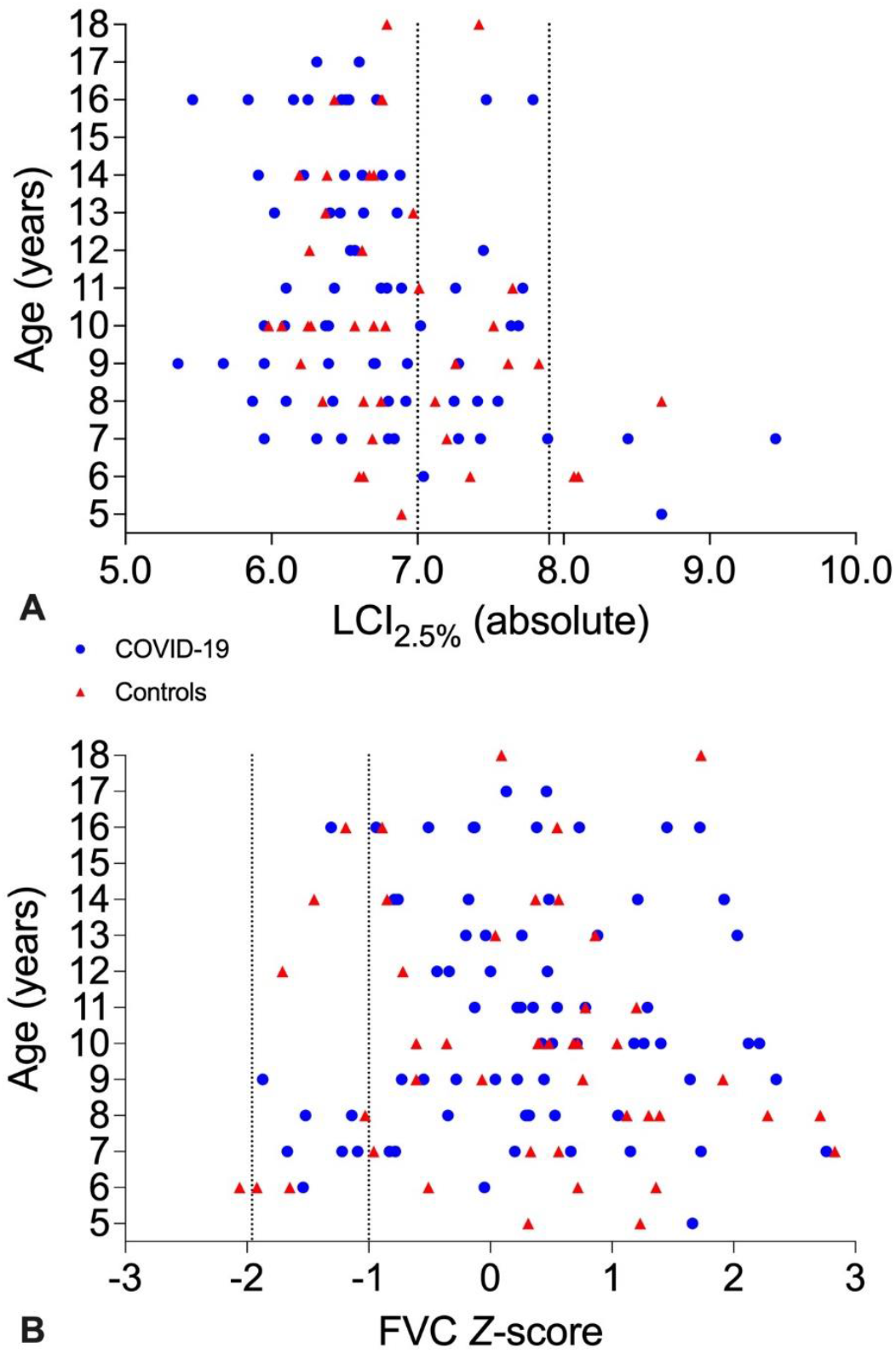
Scatter plot with **A** LCI_2.5%_ (absolute) and **B** FVC *Z*-score shown on the *x*-axis and age in years depicted on the *y*-axis. Patients after COVID-19 infection are depicted as blue dots, controls are marked as red triangles. **A** The dotted lines mark LCI_2.5%_ of 7.0 and 7.9. Values >7.0 are borderline. Values >7.9 are pathological. **B** The dotted lines mark FVC *Z*-score of –1.96 and –1. Values <–1 are borderline. Values <–1.96 are pathological. LCI_2.5%_, Lung Clearance Index at 2.5% of starting concentration; FVC, Forced Vital Capacity

Only one child with pneumonia underwent pulmonary imaging during acute COVID-19 infection. No other child or adolescent required imaging of the lungs based on medical assessment.

## DISCUSSION

Pulmonary involvement, including severe interstitial pneumonia and acute respiratory distress syndrome (ARDS), in SARS-CoV-2 infections has been the cause of high mortality and long-lasting morbidity in adults [7, 14]. In children and adolescents, severe pulmonary complications are rare [15], even though 11.1% of our patients with symptomatic SARS-CoV-2 infection described dyspnoea with acute COVID-19.

Most SARS-CoV-2 infections in our cohort, however, were asymptomatic during the acute phase. Only 35.6% of patients in our COVID-19 group stated to having had any symptoms during acute infection; respiratory symptoms were reported in 26%. In an Italian cohort with children and adolescents (*n* = 16), 87% showed symptoms at the time of diagnosis of SARS-CoV-2, 19% presenting with respiratory symptoms [8].

Long-term pulmonary symptoms persistent or emerging more than four weeks after the acute infection such as cough, dyspnoea or hyperreactive airways were described by 11.4% of our patients. This is similar to the findings of an Italian cohort study, in which 14.7% of the children had persistent respiratory problems for several months after COVID-19 infection [4]. In adults, frequency of persistent dyspnoea three months after discharge from hospital varies from 9% [16] to 54% [5]. Persistent pulmonary problems after viral infections are a well-known phenomenon in children: for example, children with respiratory syncytial virus (RSV) bronchiolitis show more wheezing episodes in the following 12 months [17] and even up to the age of six years, children with lower respiratory tract RSV infection in early childhood suffer more frequent wheeze (up to 4.3 times) than children without lower respiratory tract infection [18].

Pulmonary function in hospitalised adults is frequently impaired on discharge: Mo et al. described that 47% of their patients had reduced DLCO, total lung capacity (TLC) was decreased in 25%, 9.1% had impaired FVC and 13.6% showed reduced FEV_1_ [6]. But even three to four months after discharge, pulmonary function is still impaired in adults [5, 7, 15, 19, 20]. In a Norwegian cohort, FVC and FEV_1_ were reduced in about 10% and diffusion capacity in 24% of hospitalised adults three months after discharge [5]. Qin et al. detected significant differences in DLCO levels between discharged patients with severe and non-severe COVID-19 infection at a three-month follow-up [16]. Even after six months, lower DLCO and TLC were detected in patients with critical disease [19]. A Mexican study investigating the correlation between persistent dyspnoea and pulmonary function reported that patients with persistent dyspnoea had significantly lower FVC, FEV_1_ and DLCO values than those without [21].

Pulmonary function impairment in children after SARS-CoV-2 infection is rare, as we could show in our cohort. LCI_2.5%_ was abnormal in 4.4% of children and adolescents with COVID-19, 7% had reduced FVC and 5.3% showed impaired DLCO. No patient had pathological FEV_1_ values after SARS-CoV-2 infection. Even in children and adolescents with persistent respiratory symptoms, pulmonary function was normal in 75% of the subjects. Neither LCI_2.5%_, FEV_1_, DLCO or DLCO/VA showed any significant differences in patients and controls. These results support the preliminary findings of Bottino et al, in which seven children did not show any abnormalities in spirometry and diffusion capacity about two months after their mild SARS-CoV-2 infection [8]. In our cohort, minimal changes persisted for up to six months only in children and adolescents with more severe infection (SARS-CoV-2 or other infections). Similarly, other studies in children with viral infections, such as RSV or rhinovirus, detected acute and long-term loss of pulmonary function [17, 22, 23]. Other known risk factors for reduced pulmonary capacity such as preterm birth [24] or underlying pulmonary disease had no effects in our cohort.

We did not perform pulmonary imaging in all our patients, but only in the ones with severe pulmonary impairment. One had pneumonia at the time of initial diagnosis consistent with several studies showing radiological changes in children with acute COVID-19 [25, 15]. Fortunately, we did not detect any permanent changes on follow-up imaging.

Limitations of this single-centre design with a rather small sample size include age-dependent inconsistencies in pulmonary function performance, patient-reported symptoms and lack of information on long-term outcome of symptoms in controls. Further, our cohort did not include children and adolescents with critical respiratory involvement during acute COVID-19.

To our knowledge, our study is the first to compare pulmonary function in symptomatic and asymptomatic children and adolescents with and without evidence of SARS-CoV-2 infection; no difference between these two groups was observed. Even most patients with persistent respiratory symptoms did not show impaired lung capacity. Severity of infection proved to be the only predictor for mild pulmonary function changes. To conclude, our findings suggest that children and adolescents after SARS-CoV-2 infection do not suffer from persistent deterioration of respiratory function, including body plethysmography, multiple-breath washout and diffusion capacity testing.

Further studies with a larger, more representative cohort (including patients with critical respiratory involvement) and over a longer period are needed for better understanding of the respiratory long-term impairment after SARS-CoV-2 infection in children and adolescents. The discrepancy between subjective persistent respiratory complaints and normal pulmonary function might be caused by functional respiratory disorders, for example hyperventilation, as already described in adults [26, 27].

Thus, further studies including treadmill testing would be useful and were already initiated in our cohort.

## Data Availability

Data are available upon reasonable request.

## Acknowledgements

We are grateful to the healthcare team at the Children’s University Hospital Bochum, especially Chantal Matenar whose efforts were fundamental to this work. We would like to thank all the patients and families involved.

---

The RECORD statement – checklist of items, extended from the STROBE statement, that should be reported in observational studies using routinely collected health data.

|  | **Item No.** | **STROBE items** | **Location in manuscript where items are reported** | **RECORD items** | **Location in manuscript where items are reported** |
| --- | --- | --- | --- | --- | --- |
| **Title and abstract** |  |  |  |  |  |
|  | 1 | (a) Indicate the study's design with a commonly used term in the title or the abstract (b) Provide in the abstract an informative and balanced summary of what was done and what was found | a) and b) addressed in the abstract | RECORD 1.1: The type of data used should be specified in the title or abstract. When possible, the name of the databases used should be included.<br>RECORD 1.2: If applicable, the geographic region and timeframe within which the study took place should be reported in the title or abstract.<br>RECORD 1.3: If linkage between databases was conducted for the study, this should be clearly stated in the title or abstract. | 1.1 specified in the abstract<br>1.2 this information is included in the methods section<br>1.3 n.a. |
| **Introduction** |  |  |  |  |  |
| Background rationale | 2 | Explain the scientific background and rationale for the investigation being reported | Explained in the introduction |  |  |
| Objectives | 3 | State specific objectives, including any prespecified hypotheses | Stated in the last paragraph of the introduction |  |  |
| **Methods** |  |  |  |  |  |
| Study Design | 4 | Present key elements of study design early in the paper | Presented in methods |  |  |
| Setting | 5 | Describe the setting, locations, and relevant dates, including periods of recruitment, exposure, follow-up, and data collection | Deccribed in methods |  |  |
| Participants | 6 | (a) *Cohort study* - Give the eligibility criteria, and the sources and methods of selection of participants. Describe methods of follow-up<br>*Case-control study* - Give the eligibility criteria, and the sources and methods of case ascertainment and control selection. Give the rationale for the choice of cases and controls<br>*Cross-sectional study* - Give the eligibility criteria, and the sources and methods of selection of participants<br>(b) *Cohort study* - For matched studies, give matching criteria and number of exposed and unexposed<br>*Case-control study* - For matched studies, give matching criteria and the number of controls per case | a) cohort study: data on the items mentioned are included<br>b) cohort study: data provided | RECORD 6.1: The methods of study population selection (such as codes or algorithms used to identify subjects) should be listed in detail. If this is not possible, an explanation should be provided.<br>RECORD 6.2: Any validation studies of the codes or algorithms used to select the population should be referenced. If validation was conducted for this study and not published elsewhere, detailed methods and results should be provided.<br>RECORD 6.3: If the study involved linkage of databases, consider use of a flow diagram or other graphical display to demonstrate the data linkage process, including the number of individuals with linked data at each stage. | Described in results<br>n,a,<br>n.a. |
| Variables | 7 | Clearly define all outcomes, exposures, predictors, potential confounders, and effect | Defined in methods | RECORD 7.1: A complete list of codes and algorithms used to classify exposures, outcomes, confounders, and effect modifiers should be provided. If these | Provided in methods |
|  |  | modifiers. Give diagnostic criteria, if applicable. |  | cannot be reported, an explanation should be provided. |  |
| Data sources/<br>measurement | 8 | For each variable of interest, give sources of data and details of methods of assessment (measurement).<br>Describe comparability of assessment methods if there is more than one group | Mentioned in methods |  |  |
| Bias | 9 | Describe any efforts to address potential sources of bias | Adressed in methods |  |  |
| Study size | 10 | Explain how the study size was arrived at | Explained in methods |  |  |
| Quantitative variables | 11 | Explain how quantitative variables were handled in the analyses. If applicable, describe which groupings were chosen, and why | Explained in methods |  |  |
| Statistical methods | 12 | (a) Describe all statistical methods, including those used to control for confounding<br>(b) Describe any methods used to examine subgroups and interactions<br>(c) Explain how missing data were addressed<br>(d) *Cohort study* - If applicable, explain how loss to follow-up was addressed<br>*Case-control study* - If applicable, explain how<br>matching of cases and controls was addressed<br>*Cross-sectional study* - If applicable, describe analytical methods taking account of sampling strategy<br>(e) Describe any sensitivity analyses | a) + b) described in methods<br>c) explained in methods<br>d) cohort study, n.a.<br>e) included in the analysis |  |  |
| Data access and cleaning methods |  | .. |  | RECORD 12.1: Authors should describe the extent to which the investigators had access to the database population used to create the study population.<br>RECORD 12.2: Authors should provide information on the data cleaning methods used in the study. | n.a.<br>Described in methods |
| Linkage |  | .. |  | RECORD 12.3: State whether the study included person-level, institutional-level, or other data linkage across two or more databases. The methods of linkage and methods of linkage quality evaluation should be provided. | n.a. |
| **Results** |  |  |  |  |  |
| Participants | 13 | (a) Report the numbers of individuals at each stage of the study (*e.g.*, numbers potentially eligible, examined for eligibility, confirmed eligible, included in the study, completing follow-up, and analysed)<br>(b) Give reasons for non-participation at each stage. | a) + b) included in results | RECORD 13.1: Describe in detail the selection of the persons included in the study (*i.e.*, study population selection) including filtering based on data quality, data availability and linkage. The selection of included persons can be described in the text and/or by means of the study flow diagram. | Detailed description in results |
|  |  | (c) Consider use of a flow diagram |  |  |  |
| Descriptive data | 14 | (a) Give characteristics of study participants (*e.g.*, demographic, clinical, social) and information on exposures and potential confounders<br>(b) Indicate the number of participants with missing data for each variable of interest<br>(c) *Cohort study* - summarise follow-up time (*e.g.*, average and total amount) | a) information given in results<br>b) indicated in results<br>c) included in results |  |  |
| Outcome data | 15 | *Cohort study* - Report numbers of outcome events or summary measures over time<br>*Case-control study* - Report numbers in each exposure category, or summary measures of exposure<br>*Cross-sectional study* - Report numbers of outcome events or summary measures | Cohort study: outcome reported in results |  |  |
| Main results | 16 | (a) Give unadjusted estimates and, if applicable, confounder-adjusted estimates and their precision (*e.g.*, 95% confidence interval). Make clear which confounders were adjusted for and why they were included (b) Report category boundaries when continuous variables were categorized<br>(c) If relevant, consider translating estimates of relative risk into absolute risk for a meaningful time period | a) + b) Included in results<br>c) n.a. |  |  |
| Other analyses | 17 | Report other analyses done—e.g., analyses of subgroups and interactions, and sensitivity analyses | n.a. |  |  |
| **Discussion** |  |  |  |  |  |
| Key results | 18 | Summarise key results with reference to study objectives | Included in the paper |  |  |
| Limitations | 19 | Discuss limitations of the study, taking into account sources of potential bias or imprecision. Discuss both direction and magnitude of any potential bias | Discussed in the section | RECORD 19.1: Discuss the implications of using data that were not created or collected to answer the specific research question(s). Include discussion of misclassification bias, unmeasured confounding, missing data, and changing eligibility over time, as they pertain to the study being reported. | Included in the discussion section |
| Interpretation | 20 | Give a cautious overall interpretation of results considering objectives, limitations, multiplicity of analyses, results from similar studies, and other relevant evidence | Data interpretation evaluated in the discussion |  |  |
| Generalisability | 21 | Discuss the generalisability (external validity) of the study results | Discussed in the final paragraphs |  |  |

| Other Information |  |  |  |  |  |
| --- | --- | --- | --- | --- | --- |
| Funding | 22 | Give the source of funding and the role of the funders for the present study and, if applicable, for the original study on which the present article is based | Information provided in the funding section |  |  |
| Accessibility of protocol, raw data, and programming code |  | .. |  | RECORD 22.1: Authors should provide information on how to access any supplemental information such as the study protocol, raw data, or programming code. | n.a. |

\*Reference: Benchimol EI, Smeeth L, Guttman A, Harron K, Moher D, Petersen I, Sørensen HT, von Elm E, Langan SM, the RECORD Working Committee. The REporting of studies Conducted using Observational Routinely-collected health Data (RECORD) Statement. *PLoS Medicine* 2015; in press.
\*Checklist is protected under Creative Commons Attribution ([CC BY](https://creativecommons.org/licenses/by/4.0/)) license.

## Notes

**Funding** This study has partially been funded by the Federal Ministry of Education and Research (Bundesministerium für Bildung und Forschung [BMBF], trial registration 01KI20173).

### Competing Interest Statement

The authors have declared no competing interest.

### Clinical Trial

DRKS00022434

### Funding Statement

This study has partially been funded by the Federal Ministry of Education and Research (Bundesministerium fuer Bildung und Forschung [BMBF], trial registration 01KI20173).

### Author Declarations

Ethical approval was provided by the Ethics Committee of the Ruhr-University Bochum, Germany (No. 20-6927).

